# First GWAS on Alzheimer’s Disease in Argentina and Chile populations

**DOI:** 10.1101/2023.01.16.23284609

**Authors:** Maria Carolina Dalmasso, Itziar de Rojas, Natividad Olivar, Carolina Muchnik, Bárbara Angel, Sergio Gloger, Mariana Soledad Sanchez Abalos, M.Victoria Chacón, Rafael Aránguiz, Paulina Orellana, Carolina Cuesta, Pablo Galeano, Lorenzo Campanelli, Gisela Vanina Novack, Luis Eduardo Martinez, Nancy Medel, Julieta Lisso, Zulma Sevillano, Nicolás Irureta, Eduardo Miguel Castaño, Laura Montrreal, Michaela Thoenes, Claudia Hanses, Stefanie Heilmann-Heimbach, Claudia Kairiyama, Ines Mintz, Ivana Villella, Fabiana Rueda, Amanda Romero, Nancy Wukitsevits, Ivana Quiroga, Cristina Gona, EADB, Jean-Charles Lambert, Patricia Solis, Daniel Gustavo Politis, Carlos Alberto Mangone, Christian Gonzalez-Billault, Mercè Boada, Lluís Tàrraga, Andrea Slachevsky, Cecilia Albala, Patricio Fuentes, Silvia Kochen, Luis Ignacio Brusco, Agustín Ruiz, Laura Morelli, Alfredo Ramírez

## Abstract

**INTRODUCTION:** Genome-wide association studies (GWAS) are fundamental for identifying loci associated with diseases. However, they require replication in other ethnicities.

**METHODS:** we performed a GWAS on sporadic Alzheimer’s disease (AD) including 540 patients and 852 controls from Argentina and Chile. We explored the variants associated with AD in European GWAS from European Alzheimer’s and Dementia Biobank (EADB) and tested their genetic risk score (GRS) performance in this admixed population.

**RESULTS:** we detected *APOE4* as single genome-wide significant signal (OR=2.93[2.37-3.63], p=2.6×10^−23^), and fifteen additional suggestive signals previously undetected. Nine of the 83 variants reported by EADB in Europeans were replicated, and the AD-GRS presented similar performance in this Latin population, despite the score diminishes when the Native American ancestry rises.

**DISCUSSION:** we report the first GWAS on AD in a population from South America. It shows shared genetics that modulate AD risk between the European and the Latin American populations.

## 1. Introduction

Alzheimer’s disease (AD) is a progressive neurodegenerative disorder responsible for most dementia cases worldwide in the elderly population [1]. Although there are numerous studies on AD with the most diverse approaches, the causes and etiology of the disease remain poorly understood. Among them, genome-wide association studies (GWASs) and meta-analysis thereof have led to the identification of more than 80 genetic variants contributing to the susceptibility of AD [2]–[4]. However, the majority of these studies have been performed in European and Asian populations [5], hindering thereby their translation to populations showing different or mixed ancestries due to possible differences in the genomic structure and/or allele frequencies in each identified locus. These differences might also involve different causative variants across ancestries or allelic heterogeneity implicating, thus, alternative pathogenic mechanisms and potentially population-specific as well.

Latin American populations are diverse, not only culturally, but also in their genetic ancestry composition [6]. South American populations present a large genetic diversity in Native American and mestizo populations, between and within countries [6], [7]. This diversity is likely to have an impact on the distribution of genetic determinants of AD risk across different geographic regions. Unfortunately, systematic genetic studies for translating findings from Europeans to Latin American populations are scarce [8]–[10]. In fact, only 1.3% of individuals in the NHGRI-EBI GWAS-Catalog are Hispanic or Latin American [5]. Consequently, we report here the first GWAS on AD in a population sample from the southern cone of South America. We explored new suggestive loci and study the behavior in terms of effect size and direction of the known AD genes in a population sample from Argentina and Chile. The combined effects of these variants in a genetic risk score (GRS) can identify individuals at the highest risk of future AD [2], [3] so then, we tested the performance of the AD-GRS reported by the European Alzheimer’s and Dementia Biobank (EADB) [2] in this admixture population. Exploring different populations will likely contribute to a better understanding of the pathophysiology of AD. Importantly, understanding population-shared genetic risk factors, and the allelic and non-allelic heterogeneity of AD will translate into improved prevention and/or treatment for different populations via precision medicine.

## 2. Methods

### 2.1 Data collection

Participants in this study were obtained from multiple sources. Further sample descriptions can be found in Table 1.

**Table 1.** Descriptive characteristics of the samples across datasets.

| Cohort | Argentina<br>(n = 1018) | Chile<br>(n = 375) |
| --- | --- | --- |
| **Cases** | **416** | **123** |
| Female (%) | 66.1 | 53.7 |
| Age<sup>a</sup> (years) | 76.3 ± 6.6 | 79.6 ± 10.9 |
| *APOE*ε4<sup>b</sup> (%) | 42.8 | 50.4 |
| **Controls** | **602** | **252** |
| Female (%) | 71.2 | 69.4 |
| Age<sup>a</sup> (years) | 72.5 ± 7.5 | 81.7 ± 7.4 |
| *APOE*ε4<sup>b</sup> (%) | 19.1 | 18.7 |
| **Ancestry proportions<sup>c</sup>** |  |  |
| African | 0.05 ± 0.04 (0.00 – 0.54) | 0.04 ± 0.03 (0.00 – 0.13) |
| European | 0.73 ± 0.24 (0.00 – 0.99) | 0.57 ± 0.11 (0.19 – 0.94) |
| Native American | 0.22 ± 0.24 (0.00 – 0.97) | 0.38 ± 0.11 (0.01 – 0.77) |

<sup>a</sup>mean ± standard deviation. <sup>b</sup>Percent frequency of *APOE* ε4 allele. <sup>c</sup>Ancestry proportion, average ancestry found per individual expressed as mean ± standard deviation (minimum-maximum).

#### Argentina

The Argentine samples were recruited in the context of the Alzheimer’s Genetics in Argentina – ALZheimer ARgentina (AGA-ALZAR, https://www.gaaindata.org/partner/AGA), from the following centers: Medical Research Institute A. Lanari (C1427ARO, Buenos Aires City), Hospital de Clínicas José de San Martín (C1120AAF, Buenos Aires City), Hospital HIGA-Eva Perón (B1650NBN, General San Martín), Hospital El Cruce (B1888AAE, Florencio Varela), and several geriatric centers across Jujuy and Mendoza provinces, organized and coordinated by their respective Public Ministry of Health. The study (protocol CBFIL#22) was approved by the ethical committee (HHS IRB#00007572, IORG#006295, FWA00020769), and all participants and/or family members gave their informed consent [11]. Diagnosis of AD followed diagnostic criteria from the National Institute of Neurological and Communicative Disorders and Stroke and the Alzheimer’s disease and Related Disorders Association (NINCDS-ADRDA) [12], [13]. A total of 1542 peripheral blood or saliva samples were processed to obtain DNA using the QIAmp DNA mini kit (Qiagen); 884 controls, 133 mild cognitive impairment (MCI) and 525 AD cases. Purified DNA samples were genotyped using the Illumina Infinium Global Screening Array (GSA) v.1.0 combined to a GSA shared custom content.

#### Chile

The Chilean samples recruited correspond to patients with AD and control subjects, from different studies. Control individuals were recruited from Alexandros longitudinal study [14], that belong to 2 cohorts (SABE [15] and ALEXANDROS [16]) of community dwelling older adults of different demographic origin and socioeconomic level, mainly in the study of healthy life expectancy, free of disability and dementia. All participants were randomly selected from 18 Primary Health Care Centers and signed an informed consent on enrolment after they had received written and verbal information about the study. The ethical committee of Institute of Nutrition and Food Technology (INTA), University of Chile (Acta 23, 2012) approved the study protocol (FONDECYT nº1130947). AD patients (n=91) were recruited at Biomedica Research Group, a clinical research center performing industry sponsored international multicenter studies in Santiago. Subjects were comprehensively studied and diagnosed following the NINCDS-ADRDA [12], [13] criteria for AD. The GWAS study was approved by the Ethics Committe “Servicio de Salud Metropolitano Oriente” (SSMO). Additional AD cases and control individuals (32 AD and 20 controls) from Santiago were recruited from the GERO [17] (Geroscience Center for Brain Health and Metabolism) study at the Memory and Neuropsychiatric Center of the Hospital del Salvador and Faculty of Medicine of the University of Chile. The FONDAP GERO project n°15150012 was also approved by the Ethics Committee of the SSMO.

A total of 934 samples (n=800 DNA and n=134 frozen blood) were sent to Ace Alzheimer Center Barcelona (Barcelona, Spain) for processing. DNA was extracted from peripheral blood according to standard procedures using the Chemagic system (Perkin Elmer). For the starting DNA samples, a re-extraction protocol using the Chemagic system was also followed in order to purify the DNA samples. Only samples reaching DNA concentrations of >10 ng/µL and presenting high integrity were included for genotyping. Finally, AD cases (n=123) and controls (n=252) were randomized across sample plates to avoid batch effects. We used the Axiom 815K Spanish biobank array (Thermo Fisher) at the Spanish National Centre for Genotyping (CeGEN, Santiago de Compostela, Spain) for genotyping.

### 2.2. Quality Control and Imputation

Details on quality-control (QC) and imputation procedures are provided in previous publications [3], [18], using PLINK 2.0 [19] (www.cog-genomics.org/plink/2.0/). Briefly, individuals with low-quality samples, excess of heterozygosity, sex discrepancies, and familial relations between samples (PI-HAT > 0.1875) were excluded from the analysis. Variants with call rate below 95%, a deviation from the Hardy–Weinberg equilibrium (HWE, p < 1×10^−6^) or differential missingness between cases and controls were also removed from the analysis. To maximize genetic coverage, we performed single-nucleotide polymorphism (SNP) imputation on genome build GRCh38 using the Trans-Omics for Precision Medicine (TOPMed) imputation server [20]–[22]. Statistical power was estimated using the Genetic Power Calculator tool [23] (https://zzz.bwh.harvard.edu/gpc/cc2.html), and PowerPlot.R (https://github.com/ilarsf/gwasTools).

### 2.3. Global Ancestry Analysis

Global ancestry was estimated as described before [11]. Briefly, 446 ancestry informative markers (AIMs), specifically selected to estimate ancestry in Latin America [24], were extracted from the Latin datasets and the reference populations in 1000 Genomes (http://www.internationalgenome.org/): Caucasian (CEU, n=85), Yorubas African (YRI, n=88) and Native American [25] (NAM, n=46). From SNPs present in all populations, balanced distributed SNPs among reference populations and chromosomes, were selected to estimate ancestry (n=356). They were all merged in one PLINK v1.9 file (www.cog-genomics.org/plink/1.9/), and ancestry was predicted using ADMIXTURE v1.3.0 [26]. Plots and analysis were performed with R (www.R-project.org/).

### 2.4. Association Analysis

Logistic regression models, adjusted for age, gender and the first six ancestry principal components (PCs) were fitted using PLINK 2.0 [19] in both populations. Low imputation quality variants (R^2^ < 0.3) or rare variants (minor allele frequency (MAF) < 1%) were excluded. After study-specific variant filtering and QC procedures, we performed a fixed effects inverse-variance–weighted meta-analysis [27] with the Argentine and Chilean summary statistics for AD association. Quantile–quantile plots, Manhattan plots, and the exploration of genomic inflation factors were performed using the R package qqman [28]. Regional plots were generated with LocusZoom [29] and loci were annotated as the closest gene.

Genomic regions previously associated with AD [2] were also visualized by regional plots. *Loci* where a signal with p<0.001 was detected in the proximity to the previously reported top variant (±300 Kb), were selected for follow-up. Linkage disequilibrium (LD) estimation between these two top hits in *WDR12, INPP5D, ANKH, JAZF1, SEC61G, SORL1, FERMT2, ABCA7, ABI3 and ADAMTS1 loci* were calculated in Argentine and Chilean cohorts using PLINK 2.0 [30].

### 2.4. Genetic risk score

A weighted individual GRS was calculated based on the AD genetic variants and effect size from the recent meta-GWAS published [2] by the EADB consortium. 80 of the selected variants presented high quality in the Argentine and Chilean cohorts. The GRSs were generated by multiplying the genotype dosage of each risk allele for each variant by its respective weight and then summing across all variants. GRS association with AD cases were tested by a logistic regression model adjusted by 4PCs in each cohort. Influence of NAM ancestry over GRS was estimated by a linear regression model adjusted by sex, age and phenotype (control=0, case=1) in pooled Argentine and Chilean samples. The linear model was plotted separated for cases and controls to test interaction between NAM ancestry and disease. In addition, pooled samples were split in quintiles using NAM ancestry proportion. Differences in GRS values among quintiles were assessed by ANOVA followed by Tukey post-hoc test, and GRS association in each quintile was tested using the same logistic regression model described above. Differences in frequency between the most European (quintile 1 and 2) and the most NAM individuals (quintile 4 and 5) were estimated by a logistic regression model of ancestry (mostEUR=0, mostNAM=1) vs the 80 SNPs, adjusted by phenotype, sex, and age; p-values were Bonferroni corrected. All analyses were performed with R (www.R-project.org/).

## 3. Results

### 3.1. Population admixture in Argentinian and Chilean samples

Genome-wide genotyped data was generated in two samples from the southern cone of Latin America (Table 1), Argentina (n=1018) and Chile (n=375). We first explored the ancestry admixture of both populations (Figure 1). While the admixture of Chilean participants is more homogenous, with 75% of the samples showing 30-50% NAM ancestry, the Argentinian samples showed more diverse admixture along the NAM and EUR axis, with 32% of individuals having >30% NAM ancestry (Figure 1 and Supplementary Fig.1). Beside differences in recruitments between the Chilean (only one city, Santiago) and the Argentinian samples (different cities across the country), dissimilar migratory flows and policies between countries may explain these differences in ancestry proportions. Importantly, this admixture distribution is similar in cases and controls in both cohorts (Supplementary Fig.1).

**Figure 1.**
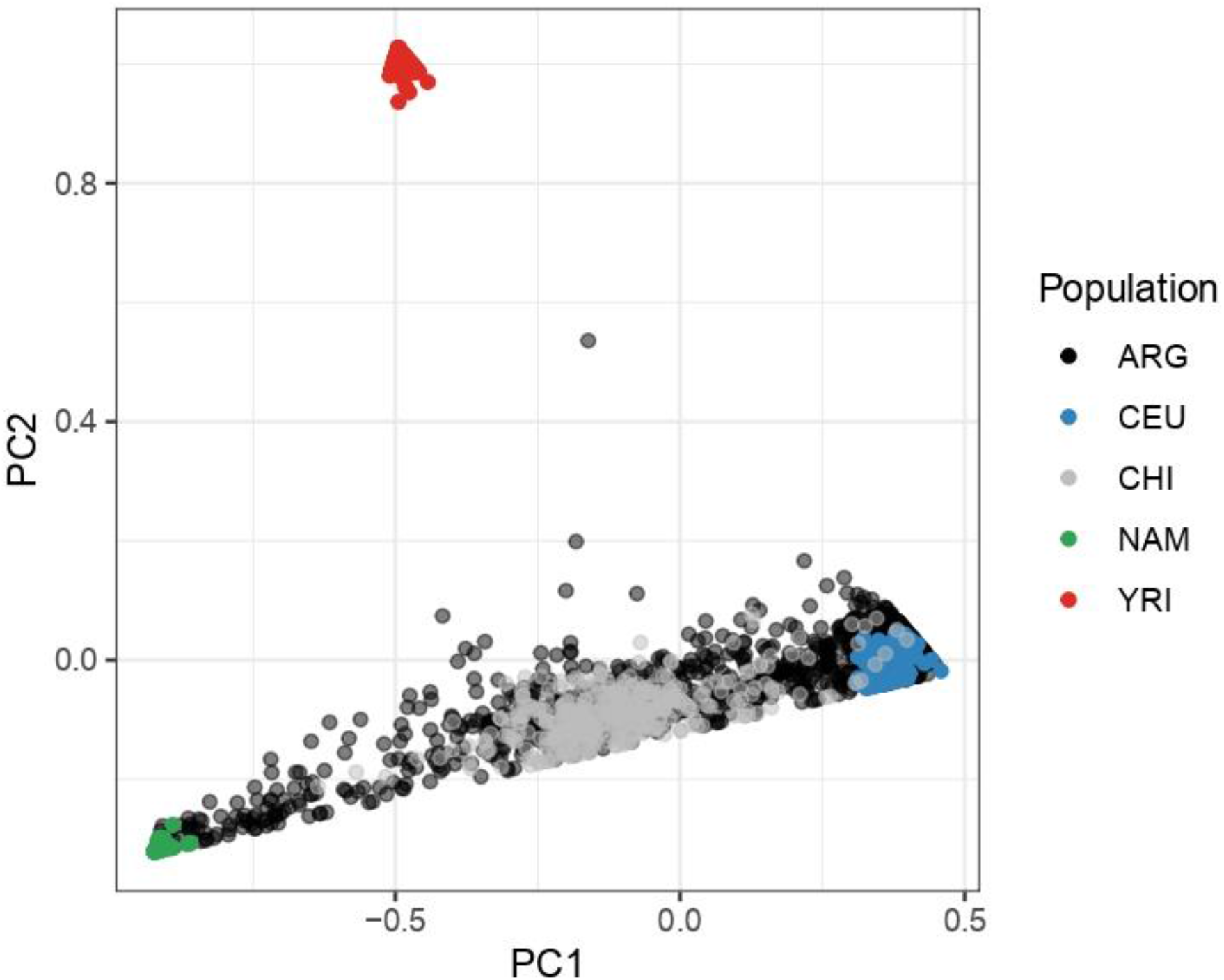
Ancestry analysis of the Argentinian and Chilean populations. Principal component analysis (PCA) of ancestry results for the Argentinian sample (ARG, black) and the Chilean sample (CHI, grey). Ancestral populations are Caucasians (CEU, blue), Yoruba (YRI, red) and Native Americans (NAM, green).

### 3.2 GWAS meta-analysis

GWAS was performed on each cohort separately and meta-analyzed as described in Materials and Methods. The combined sample size was 539 patients with AD dementia and 854 controls. Four principal components corrected inflation (λ=1.01, Supplementary Fig.2). As expected for a sample size with limited statistical power (Supplementary Fig.3), only the *APOE* locus showed an association with the risk of AD reaching genome-wide significance (rs429358-*APOE*ε4 OR=2.93[2.37-3.63], p=2.6×10^−23^; *APOE*ε2-rs7412 OR=0.53[0.34-0.84], p=6.3×10^−3^, Supplementary Fig.2). Fifteen loci reached a suggestive p-value, i.e., 5×10^−8^<p<1×10^−5^ (Table 2). However, neither of these loci was previously reported in association with AD risk in case-control GWASs nor showed nominal significance (p<0.05) in the EADB stage I [2]–[4] (Supplementary Fig.2 and Supplementary Table 1). Noteworthy, among these suggestive signals, those at *MRPL50P1* and *GPX4* deserve further mention (Table 2). At *MRPL50P1* locus, a suggestive association (rs13002275) was previously reported in a GWAS of hippocampal volume in AD [31]. This variant is in LD with our top signal rs36039096 at the same locus, with a D’=0.91 and low r^2^=0.14 due to the difference in allele frequency (MAF_rs13002275_=0.39 vs MAF_rs36039096_=0.21 in Ad Mixed American (AMR, https://www.ncbi.nlm.nih.gov/snp/ and https://ldlink.nci.nih.gov/). On the other hand, the suggestive signal in *GPX4* is located close (52.6 Kb) to the known AD locus *ABCA7*. However, the top SNP signal in our study (rs8103283) does not show LD with the top signal described for *ABCA7* in European ancestry (D’=0.19, r^2^=0.02 in AMR, https://ldlink.nci.nih.gov/). Besides, e-QTL analysis (https://gtexportal.org/) showed that rs8103283 is modulating the expression of *GPX4, POLR2E*, and *SBNO2* expression but not of *ABCA7*. Hence, *GPX4* might represent an independent signal which needs further confirmation in larger samples.

**Table 2.**
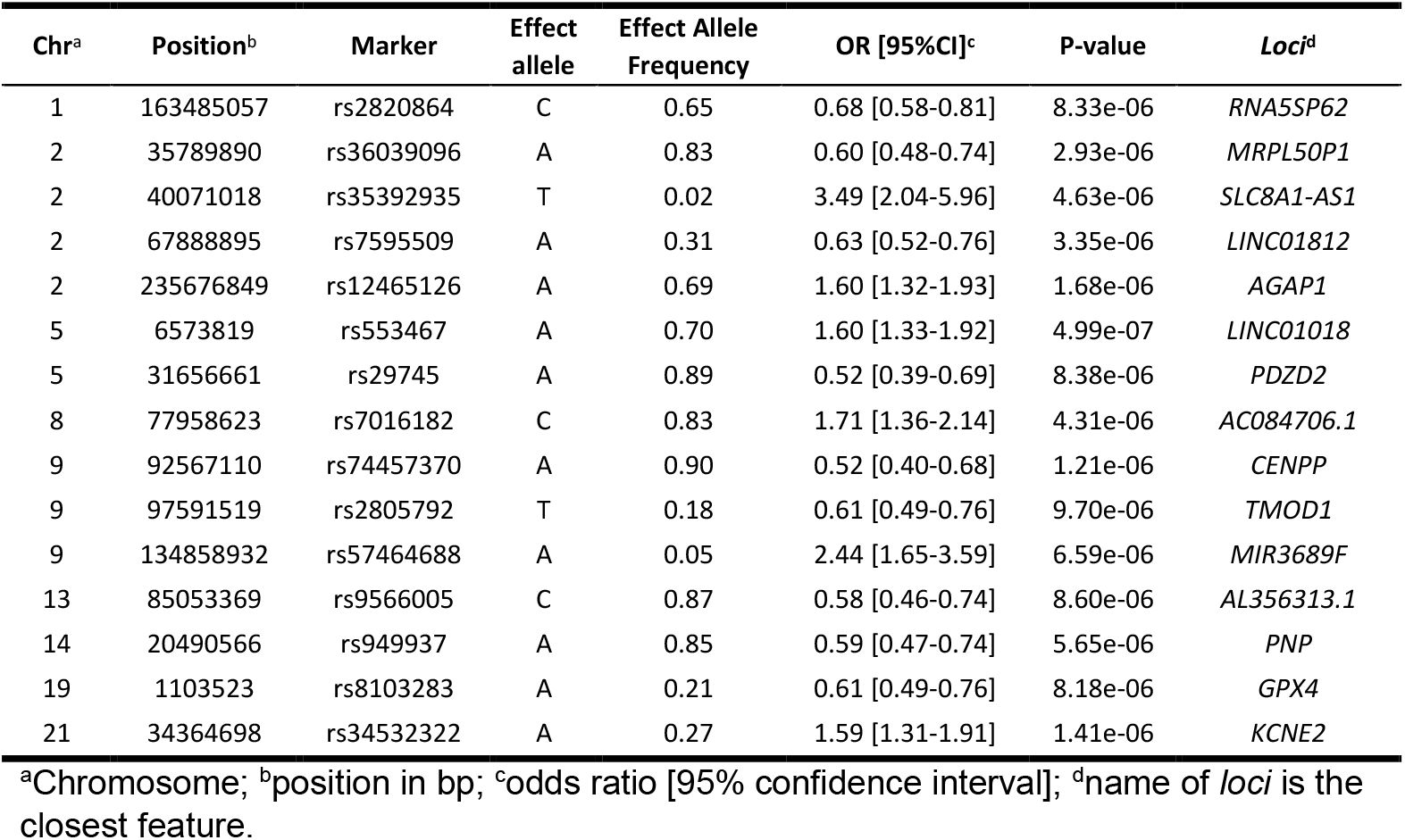
Suggestive SNPs in Argentina–Chile meta-analysis.

### 3.3 EADB hits in the Latin population meta-analysis

Next, we explore whether the genetic loci previously associated with AD risk [2] in European ancestry translate to the Latin genetic admixture included in our study. First, we investigated the 83 sentinel signals reported by EADB in our meta-analysis. Two of them were excluded from the Chilean dataset (*PLCG2* p.P522R, rs72824905 and *ABI3* p.S209F, rs616338) [2], [32], and one from the Argentinian dataset (rs7157106 at *IGH gene cluster*) due to bad quality; their associations in the remaining population are reported (Figure 2 and Supplementary Table 2). Formal replication was observed for nine variants, i.e., an identical SNP displaying the same effect direction and nominal statistical significance (p<0.05, Figure 2), representing a translation of these signals in the Latin population. It is noteworthy that in previous studies, rs17020490 at the *PRKD3* locus and rs10131280 at the *IGH-gene-cluster locus*, reached GWAS-significance in the final stages [2], [3]. Hence, we provide nominal and independent replication confirming both loci.

**Figure 2.**
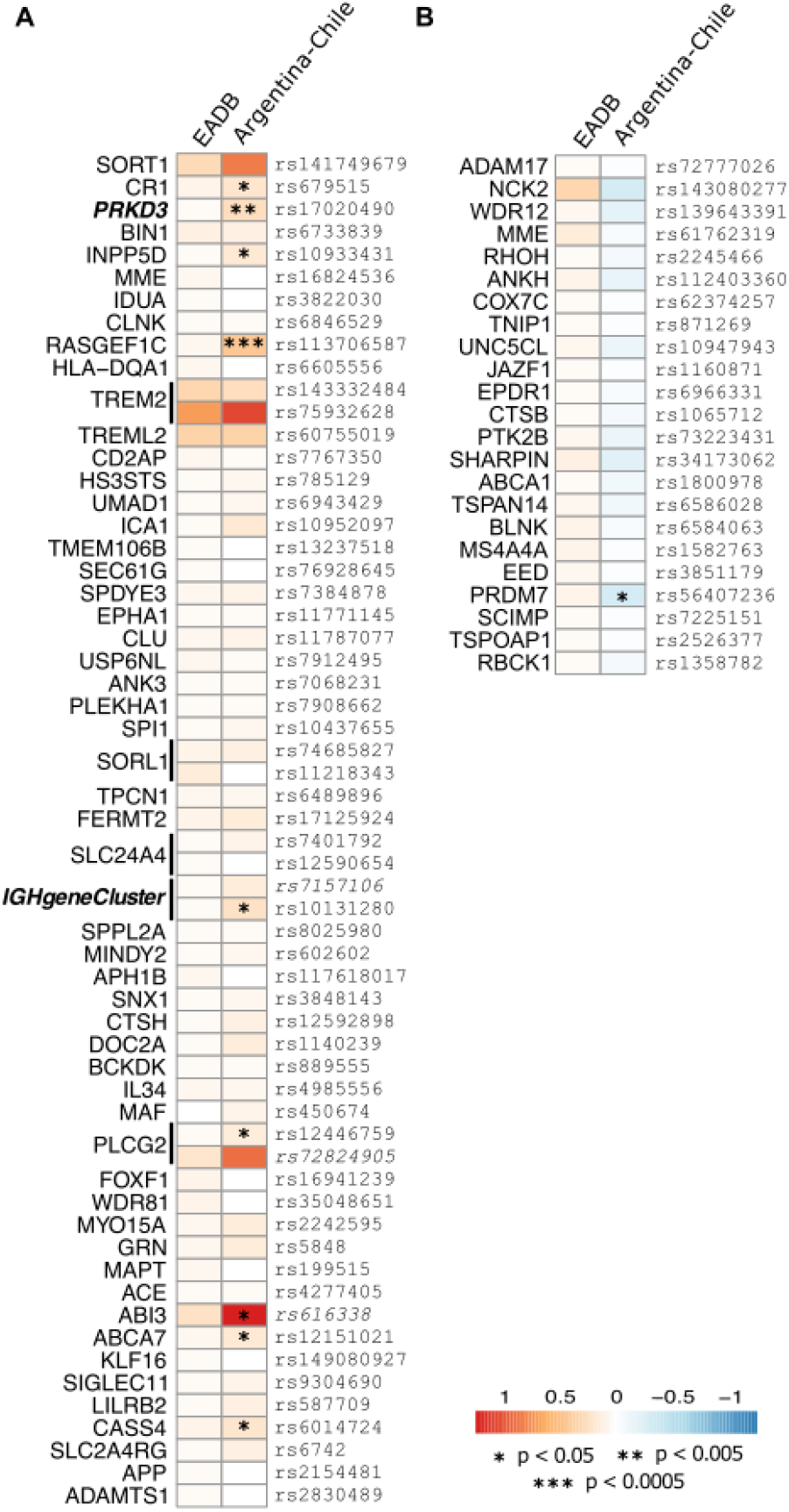
Heatmap comparison between the genome wide association SNPs in the EADB and Argentina-Chile meta-analysis. **A**. SNPs with the same direction of the effect in both meta-analyses. **B**. SNPs with the opposite direction of the effect in the Latin samples. Betas of each risk allele in EADB are in the left column and in the Argentina-Chile meta-analysis in the right column. SNP names (rs) are at the right and loci names at the left of the columns. Positive beta values are red and negative beta values in blue. Asterix represents nominal p-value (*, p<0.05; **, p<0.01; ***, p<0.001). SNP names in italic are present only in the Argentinian or Chilean sample.

Then, we analyzed sentinel SNPs in their surrounding genome region. The inspection of regional plots showed nine risk loci signal reaching a p-value∼5×10^−4^. LD estimations revealed that only the signal in *WDR12* showed LD with the top hit in EADB (Supplementary Table 3). Another signal (rs12718937) was found within the *SEC61G* locus [2], in close proximity to the gene *EGFR* (OR=0.68[0.57-0.81], p=1.70×10^−5^). Restrained LD between rs12718937 and the EADB sentinel variant rs76928645 was observed (Supplementary Table 3). Noteworthy both SNPs, seems to modulate the expression of *EGFR* as seen by e-QTL analysis (https://gtexportal.org/). Thus, our results provide independent support for *EGFR* as the most interesting candidate gene for this locus.

A known locus that deserves mention is the signal found in the *NDUFAF6* locus (rs2044899: OR=0.67[0.55-0.80], p=1.90×10^−5^). This locus was first reported in a gene-based analysis [33] and further confirmed in GWAS meta-analysis [2], [3], [18]. However, this locus was not confirmed in the last meta-analysis reported by the EADB consortium because the signal was not replicated in the stage II [2]. The signal found in our analysis (rs2044899) shows restrained LD with the signals described previously in European ancestry [2], [18] (Supplementary Table 3), suggesting that all SNPs contribute to the same susceptibility signal of AD. Further studies are necessary to confirm our observation.

### 3.4 EADB genetic risk score performance in Latin Population

Finally, we sought to explore whether the GRS reported by the EADB [2] consortium can classify cases and controls accurately in the Latin American population. To compute the GRS in our sample, we included the 80 SNPs that passed quality controls in both, the Argentinian and Chilean datasets, with the effect sizes reported in European ancestry (Supplementary Table 4). GRS values were normally distributed and logistic regression analysis revealed an association with AD in both Argentine (GRS_mean_=50.4, GRS_range_[40.1-61.8], OR=1.06, p=7.4×10^−4^) and Chilean (GRS_mean_=49.5, GRS_range_[39.3-60.9], OR=1.16, p=1.6×10^−6^) populations.

Since the Latin American population analyzed here are genetic admixtures, we investigated whether the NAM ancestry was affecting the GRS values and/or its association with the disease. A linear regression model showed that the proportion of NAM ancestry is indeed modulating the GRS values (Effect size (β)=-4.84, p<2×10^−16^), without interacting with the disease (Supplementary Fig.4). To explore this observation in detail, we split the Latin population in quintiles depending on NAM ancestry proportion (Figure 3). Quintiles 1 to 3, containing larger proportion of Caucasian ancestry individuals, showed GRS values not significantly different among them. Conversely, quintiles 4 and 5, containing higher proportion of NAM samples, showed GRS values significantly different between them, and smaller than those observed in quintile 1-3 (p<0.001). Noteworthy, while the GRS mean value decreases as the NAM ancestry proportion increases, the GRS association with AD remains similar in each quintile. In fact, the effect size for the GRS association is the same in quintile 1 than quintile 5 (Figure 3).

**Figure 3.**
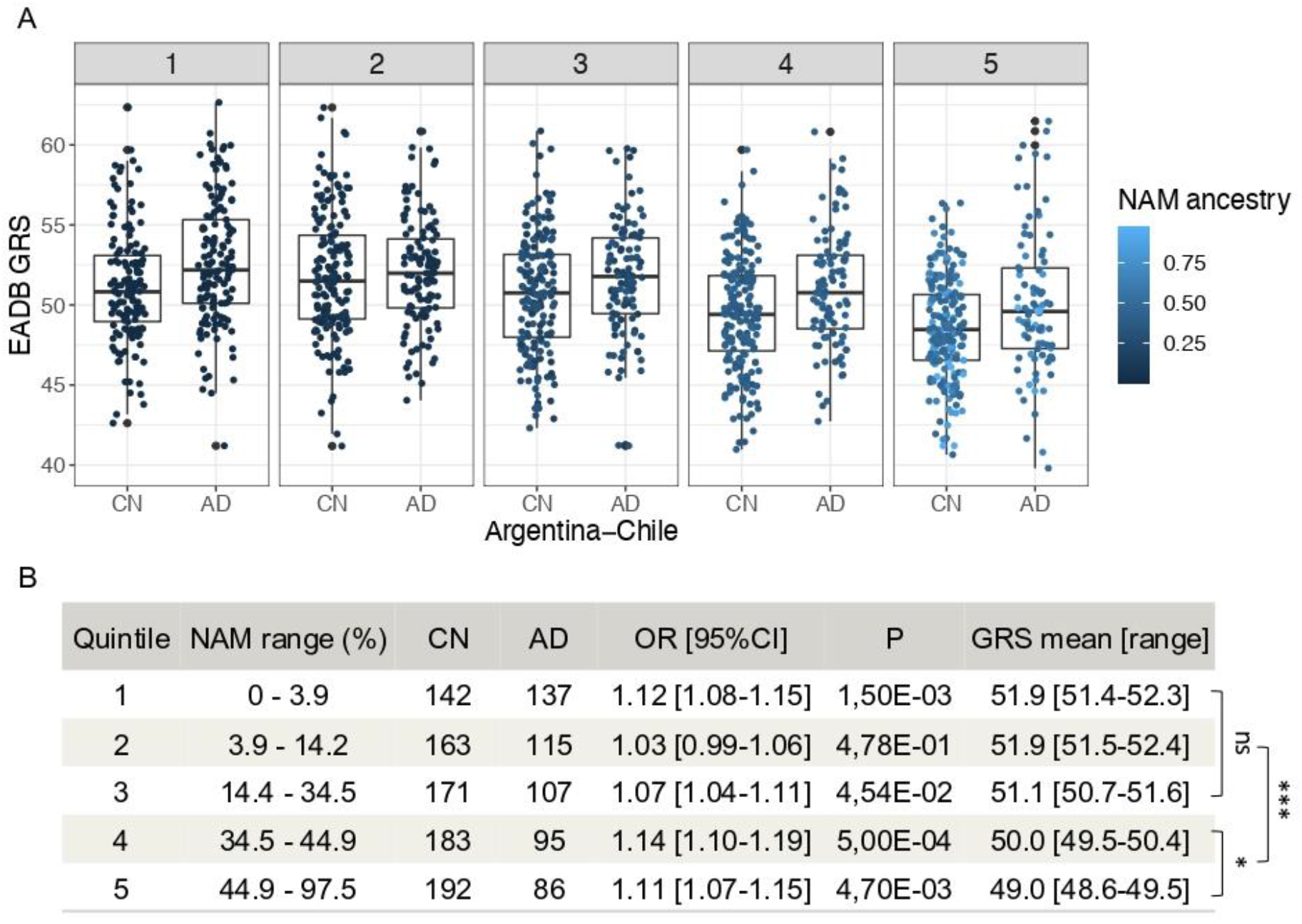
GRS performance and its association with NAM ancestry. GRSs of the samples from Argentina and Chile were split in 5 groups (quintiles) depending on their proportion of NAM ancestry. **A**. Boxplot of GRSs in cases (AD) and cognitively normal individuals (CN) present in each quintile (1 to 5). Dot color represents the degree of NAM ancestry of the sample, the lighter the higher is the proportion of NAM ancestry. **B**. Quantitative information of the quintiles. NAM range (%), proportion of NAM ancestry range; CN, number of control samples; AD, number of cases samples; OR [95%CI], GRS effect expressed as odds ratio and 95% confidence interval; P, OR associated p-value; GRS mean [range], mean value of GRSs and its respective range. At the right of the table, differences among GRS values estimated by two-way ANOVA (Tukey’s multiple comparisons test) are represented; ns, not significant; *, p<0.05; ***, p<0.001.

Differences in GRS values depends on the frequency of risk alleles in the population analyzed. Consequently, the differences observed in the GRS values in samples with higher proportion of NAM ancestry may be explained by differences in the risk allele frequency between European and NAM ancestries. To test this hypothesis, we combined quintiles 1 and 2 in one group (mostEUR) and quintiles 4 and 5 in the mostNAM group, and compare risk alleles frequencies for each of the 80 SNPs included in the GRS between groups. This comparison showed that allele frequency between both groups was significantly different (p_Bonferroni_<0.05) in 38 SNPs, of which 24 showed a lower frequency and 14 a higher frequency in the mostNAM group (Supplementary Tables 5 and 6).

## 4. Discussion

Understanding the genetics of AD is one of the best ways of improving our knowledge of the underlying pathophysiological processes. In this regard, GWAS have been pivotal for the identification of genomic regions associated with the disease. Unfortunately, large international initiatives have focused their research on European ancestry limiting thereby the generalizability of genetic findings across populations with different ancestries [5], [34]. Herein, admixture populations living in Latin America represent still a major gap for genetic research [10]. To begin filling this gap, we present here the kickoff study to elucidate AD genetics in the understudied South American population. We carried out the first AD GWAS using 1,393 samples from Argentina and Chile, generating the first GWAS summary statistics accessible for these southern populations.

While our study lacks the statistical power for claiming new population-specific signals, it is suitable for replication and translation of previously validated loci. Consequently, we provide here an extensive analysis of the main associations reported in European AD GWAS [2]–[4]. We confirmed our previous observation for the *APOE* locus, and provide independent validation for nine of the 83 SNPs tested, evidencing that they can be translated from Europeans to Latin population. Among these translated signals, we provide the first independent replication for rs10131280-*IGH-gene-cluster* loci, and rs17020490-*PRKD3*. In our study, we confirm that both loci contribute to AD susceptibility in populations other than the European’s. Additionally, we validate a common variant in the *PLCG2* locus, which together with our previous observation [11] reinforces the contribution of this locus to the susceptibility of AD in the Latin population.

We also observed nine additional signals surrounding confirmed AD loci and showing a significance of non-adjusted p<5×10^−04^. This observation might suggest the presence of allelic heterogeneity in some AD loci. However, these novel hits will require independent replication in future studies. Conversely, *WDR12* showed LD between the top hit identified in the Latin American population and the signal identified in European ancestry. Thus, *WDR12* can also be considered a locus that contributes to AD risk in the Latin American population which, however, requires further validation in larger samples from this ancestry. Likewise, the SNP rs12718937, close to the *EGFR* gene within the *SEC61G* locus, showed a nominal significance in our GWAS. Interestingly, gene prioritization in the *SEC61G* locus in Europeans suggests that *EGFR* was the only risk gene [2]. Our results provide further support for *EGFR* as the risk gene for this locus. In this case, our GWAS in a different ethnic background is helping to name the correct candidate gene within a locus.

Despite these encouraging results, we observed a low replication rate in our target population that it is in part due to the limited sample size, but also due to the presence of admixture, mainly between Amerindians and Europeans in our series. This latter issue probably caused signals to differ from those identified in Caucasians underscoring the importance of translation analysis of associated variants in different ethnicities. Given the increasing population diversity observed in countries all over the world, understanding population-shared and -specific risk factors of AD will translate into improved and specific prevention and/or treatment. To date, research has shown that GRS generated from European ancestry GWAS works more accurately in Europeans than in non-Europeans [34], [35]. In our hands, the AD-GRS developed in Europeans [2] presented similar performance in the Argentinean and Chilean populations (OR=1.09, p=3.14×10^−8^) as in European/Spanish population (GR@ACE[3], OR=1.095, p=9.63×10^−88^), independently of the degree in NAM ancestry present in the target. This means that this GRS could be generalized also to Hispanics/Latinos, as it was observed for other phenotypes [36], [37]. This can be explained because the admixture found in Argentinians and Chileans includes different proportions of European ancestry. On the other hand, GRS trans-ethnic performance also seems to depend on the sample size of the discovery GWAS. Thus, it is also possible that this GRS performed well in our Latin American sample because the EADB GWAS [2] was large enough (>500K individuals) to calculate accurate effect sizes to be used as SNP weights.

Interestingly, GRS values decreases as the NAM ancestry proportion increases. While this observation could be a real difference between the risk of AD in Europeans and Latins, this reduced GRS values seems more likely caused by incorrect variant selection and/or genetic effect used in the GRS for the target population. In other words, the genetic variants included in the GRS might be explaining less of the genetic driving AD in this ethnic admixture. Supporting this hypothesis, we observed that several SNPs included in the GRS showed significantly different risk allele frequency between NAM and European ancestry. This may complicate direct practical use of GRS score, and/or set-up a pathological predictive threshold. Further studies are needed to understand how to overcome this difficulty.

Our work has some limitations, it does not have the statistical power for discovery GWAS and/or validation of low frequency allelic associations, so we might have missed some genuine signals linked to the NAM ancestry, as well as true associations. In addition, this work might not be representative enough of the allelic variability present in Argentina and Chile, because of their vast territories and the limited number of recruitment centers included in the study. Still, our strength is to start generating genetic information on AD in the southern cone of South America, and start identifying trans-ethnic signals, which contributes to diversity studies.

## 5. Conclusions

In conclusion, we provide here the first of a series of AD GWAS to come involving population originating from countries from Latin America. Our analysis clearly showed shared genetics between the European and the Latin American populations modulating the risk of AD. However, several of these loci carry probably different genetic risk variants that should be added when constructing a GRS in Native American ancestry. Furthermore, a larger initiative is now starting to increase the sample size studied in Latin America which will lead to definition of population specific estimators for the risk conferred by each variant included in the GRS. Finally, genetic research in the Latin American population will help improving the definition of personalized risk profiles informing on the individual risk for progressing to dementia. This will likely improve our possibilities for early personalized intervention to prevent or postpone dementia.

## Data Availability

All data produced in the present study are available upon reasonable request to the authors.

## Author Contributions

ARa, LM and ARu designed, conceptualized and supervised the study, interpreted the data and revised the manuscript. MCD and IdR contributed to data acquisition, the analysis, interpreted the data and co-wrote the manuscript. Data generation and sample contribution - **Argentina**: MCD, NO, CM, PG, LC, MEC, CL, CF, MS, MF, GJ, MSSA, LEM, NM, JL, ZS, MIB, FDG, EMC, CK, JSA, HS, FJ, CAM, PS, DGP, SK, LI, LM, ARa; **Chile**: SG, BA, VC, PO, PF, ARu and IdR. All authors critically revised the manuscript for important intellectual content and approved the final manuscript.

## Data Availability Statement

The summary statistics of the meta-analysis are available to the corresponding author upon request.

## Acknowledgments

IdR. is supported by national grant from the Instituto de Salud Carlos III (ISCIII) FI20/00215. LC is supported by a doctoral fellowship of CONICET (Argentina). MCD, PG, EMC, SK and LM are members of the Research Career of CONICET (Argentina). SK, PS and MCD are supported by the Agencia Nacional de Promoción Científica y Tecnológica (PUE060, PICTO-2021-UCTH00005, PICTO-2021-UCTH00006). ARu and MB receive support from the European Union / EFPIA Innovative Medicines Initiative joint undertaking ADAPTED and MOPEAD projects (grant numbers 115975 and 115985, respectively). MB and ARu are also supported by national grants PI13/02434, PI16/01861, PI17/01474, PI19/01240, PI19/01301 and PI22/01403. Acción Estratégica en Salud is integrated into the Spanish National R+D+I Plan and funded by ISCIII –Subdirección General de Evaluación and the Fondo Europeo de Desarrollo Regional (FEDER–‘Una manera de hacer Europa’). ARu is also funded by JPco-fuND-2 “Multinational research projects on Personalized Medicine for Neurodegenerative Diseases,” PREADAPT project (ISCIII grant: AC19/00097), and EURONANOMED III Joint Transnational call for proposals (2017) for European Innovative Research & Technological Development Projects in Nanomedicine (ISCIII grant: AC17/00100). Ace Alzheimer Center Barcelona researchers research receives support from Roche, Janssen, Life Molecular Imaging, Araclon Biotech, Alkahest, Laboratorio de Análisis Echevarne, and IrsiCaixa. AS, PO and CG-B are supported by grant ANID/FONDAP (ID15150012). JCL is supported by a grant (EADB) from the EU Joint Programme – Neurodegenerative Disease Research. INSERM UMR1167 is also funded by the INSERM, Institut Pasteur de Lille, Lille Métropole Communauté Urbaine and French government’s LABEX DISTALZ program (development of innovative strategies for a transdisciplinary approach to AD).

## Supplementary Material

### Supplementary Figures

**Supplementary Figure 1.**
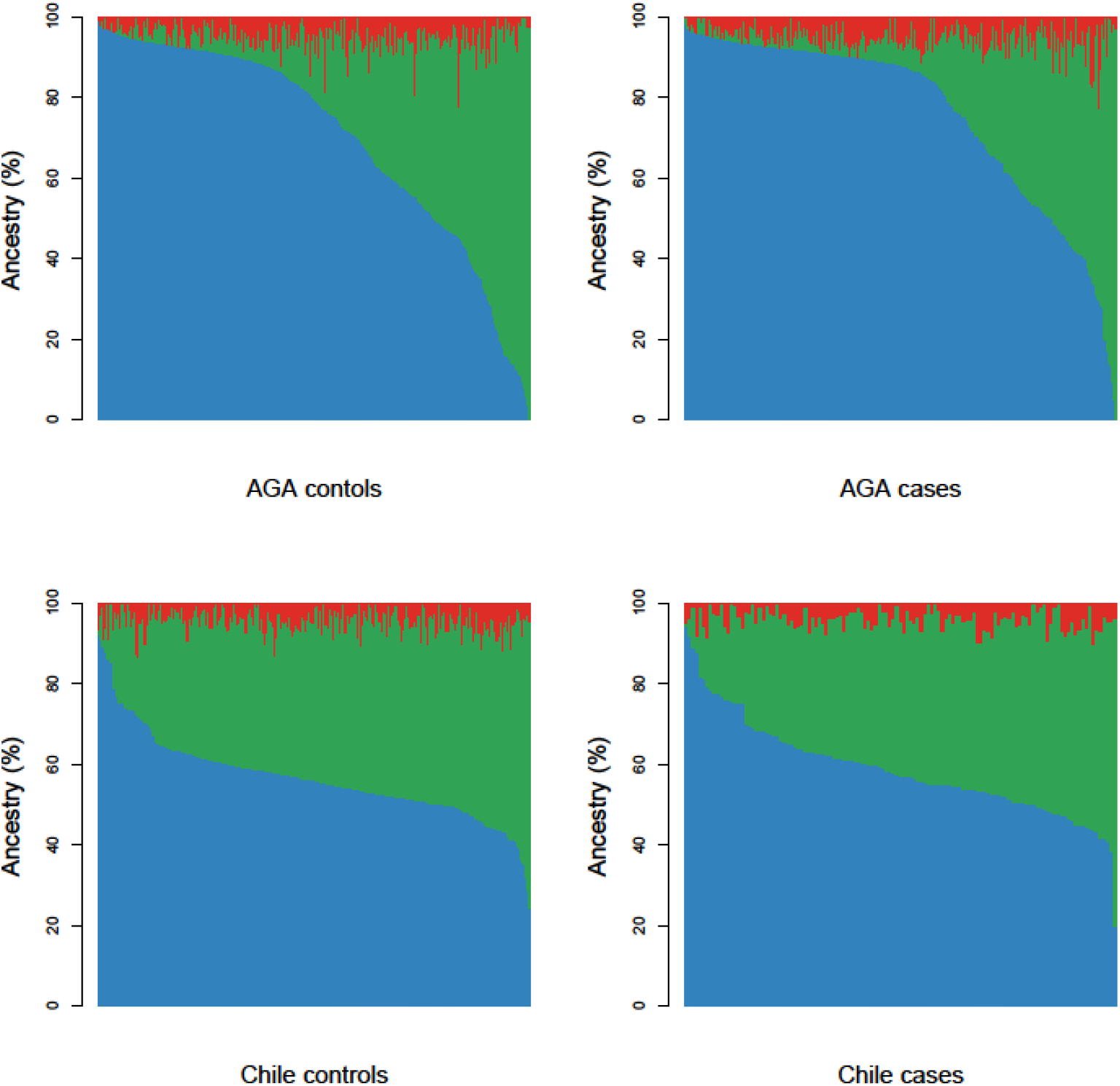
Ancestry analysis of populations. Bar-plots of each sample (x-axis) *versus* their respective percent of Caucasian (CEU, blue), African (YRI, red), and Native American (NAM, green) ancestry (y-axis). **A**. Argentinian cognitively normal samples. **B**. Argentinian AD samples. **C**. Chilean cognitively normal samples. **D**. Chilean AD samples.

**Supplementary Figure 2.**
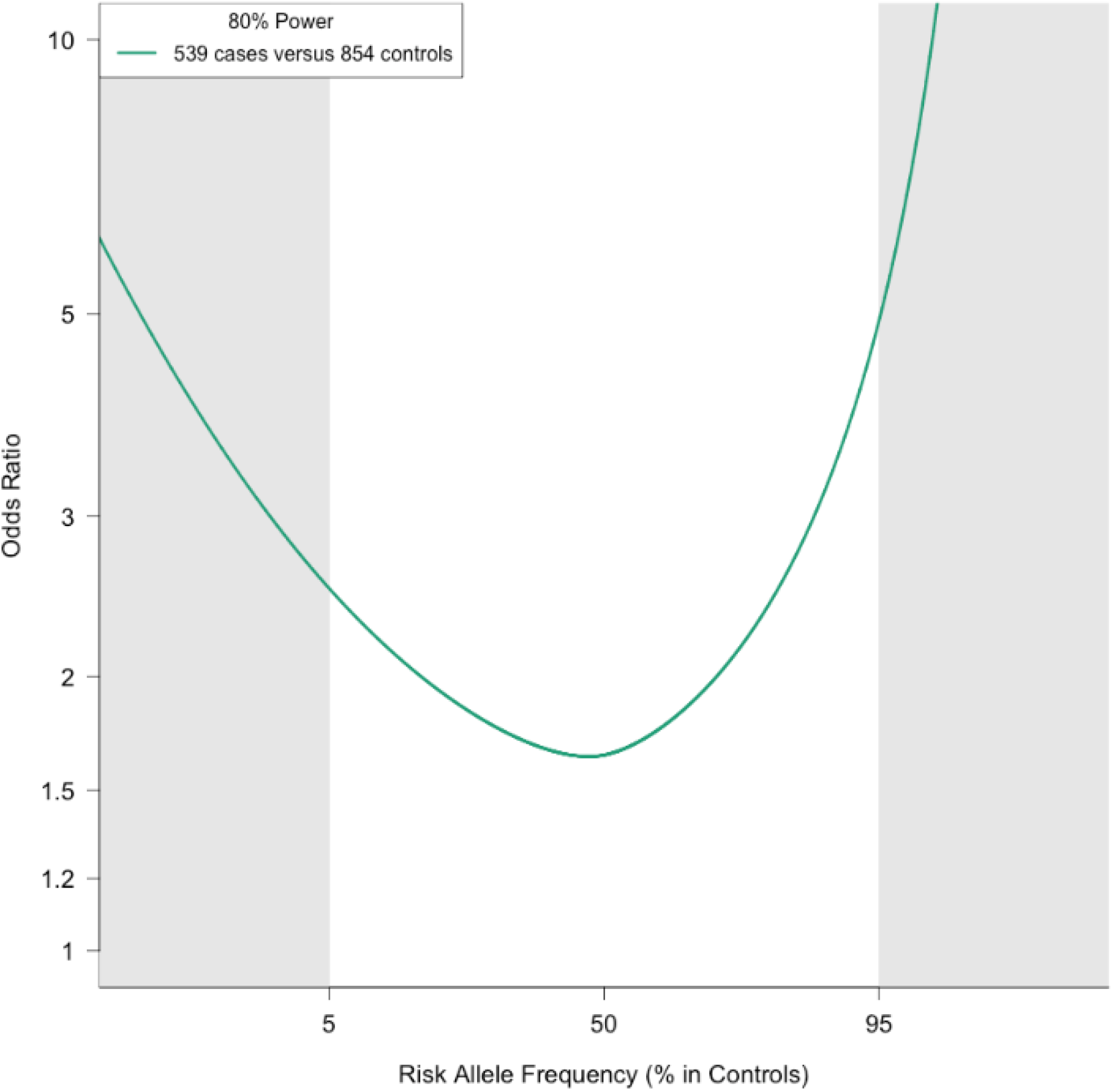
Power calculation for the meta-analysis in Latin American population. Line plot represents the Argentina-Chile meta-analysis (green line) 80% power for SNP detection, depending on the risk allele frequency (x-axis) and the odds ratio (y-axis).

**Supplementary Figure 3.**
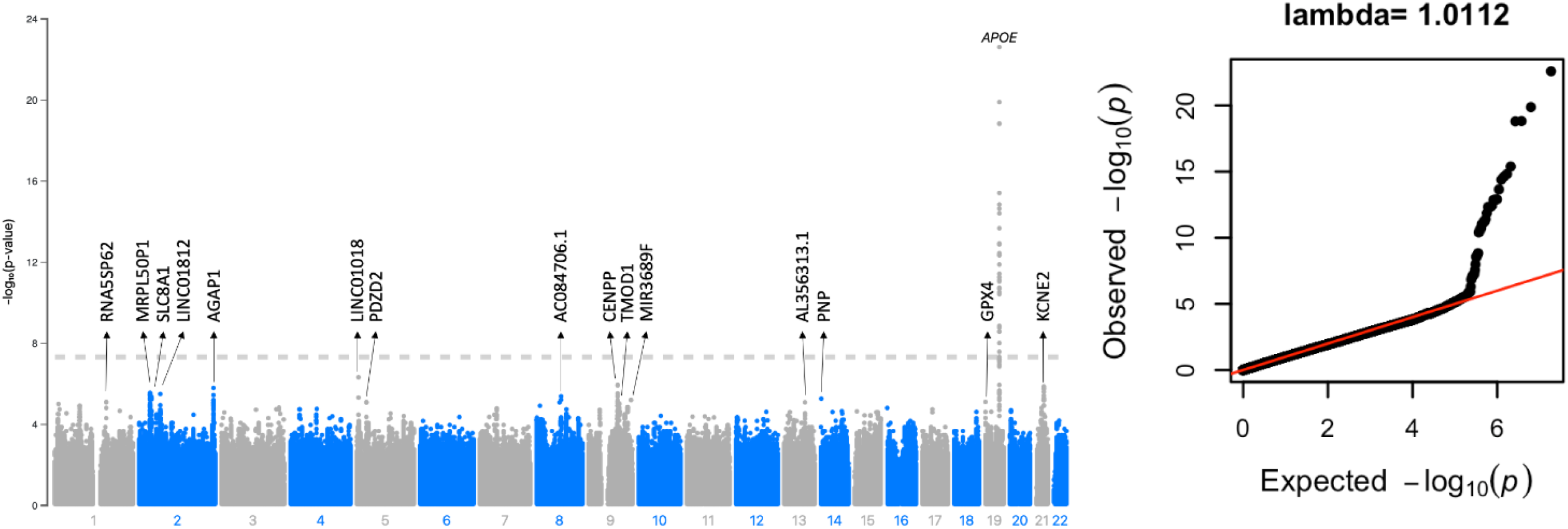
Manhattan Plot and QQplot for the Argentina-Chile meta-analysis. a) Manhattan plot. The dotted line represents the genome-wide significance level (P=5×10^−8^). *Loci* designated with arrows have suggestive significance level (P=1×10^−5^). Annotation is based on the closest gene. **B) QQplot**. Genomic inflation factor (lambda) was 1.011 when restricted to variants with minor allele frequency (MAF) above 1% (9,364,706 SNPs included).

**Supplementary Figure 4.**
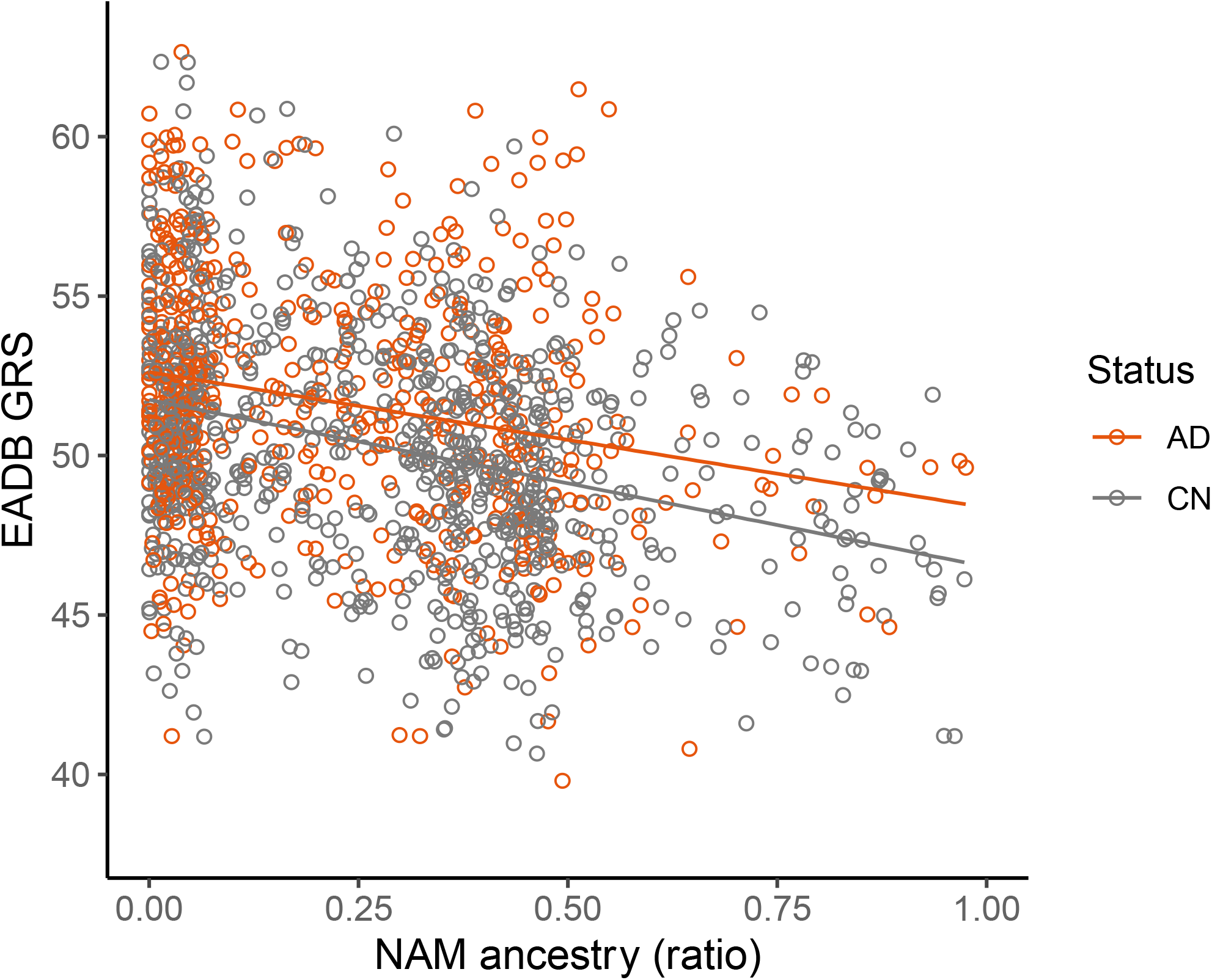
Graphical representation of linear regression model of the GRS vs NAM Ancestry. Linear regression model comparing NAM ancestry (x-axis) and GRS values (y-axis). AD-cases are in orange and controls (CN) in gray. Lines and shadings represent the linear fits and 95% confidence interval, respectively.

### Supplementary Tables

**Supplementary Table 1.**
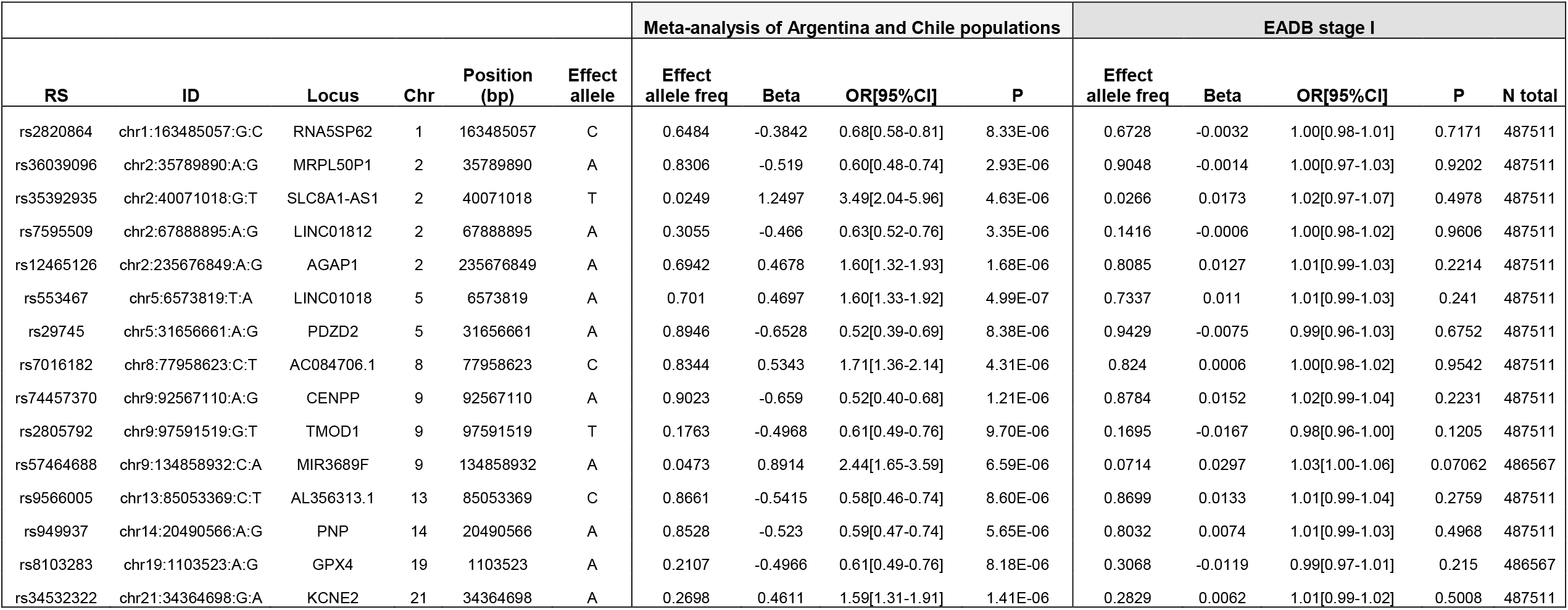
Suggestive SNPs in Argentina–Chile meta-analysis compared to EADB stage I. Abbreviations: EADB, European Alzheimer’s and Dementia Biobank; Chr, Chromosome; Freq, Frequency; OR [95% CI], Odds Ratio [95% confidence interval]; P, p-value; N total, total number of alleles.

***[See supplementary material (*.*xlsx file)]***

***Supplementary Table 2*. Replication of the SNPs associated with Alzheimer’s disease selected from the European Alzheimer’s and Dementia Biobank in Argentina and Chile populations**. Results obtained with an association and fixed-effects inverse-variance-weighted meta-analysis. Effect reported by minor allele. P-value for significance <5×10^−08^. P-value for suggestive associations <1×10^−07^. Highlighted in red p-value for replicated loci (p<0.05). OR: Odds Ratio, MAF: Minor allele frequency, Rsq: Imputation quality.

**Supplementary Table 3.** Loci detected in the meta-analysis of Latin American populations in the same regions than EADB. Abbreviations: Chr, Chromosome. ^a^Locus GW in EADB Stage I, but not replicated in the Stage II. ^b^Locus reported in a GWAS meta-analysis S. Moreno-Grau et al. 2019. Distance: base pairs between the rs in the meta-analysis of Latin American populations and EADB-rs. LD based on https://ldlink.nci.nih.gov/

| Locus | Chr | EADB Top hit |  |  | Argentina and Chile meta-analysis top signal in same locus |  |  |  |  | Distance (Kb) | Argentina |  | Chile |  |
| --- | --- | --- | --- | --- | --- | --- | --- | --- | --- | --- | --- | --- | --- | --- |
|  |  | ID | rs | Position (bp) | ID | rs | Position (bp) | Beta | P-value |  | r2 | D' | r2 | D' |
| WDR12 | 2 | chr2:202878716:TC:T | rs139643391 | 202878716 | chr2:203051767:G:A | rs11683327 | 203051767 | -0.309 | 3.32E-04 | 173.1 | 1.77E-02 | 0.403 | 7.51E-02 | 1 |
| INPP5D | 2 | chr2:233117202:G:C | rs10933431 | 233117202 | chr2:233315997:G:C | rs18115815 | 233315997 | -0.589 | 1.02E-04 | 198.8 | 7.23E-03 | 0.165 | 1.63E-02 | 0.205 |
| ANKH | 5 | chr5:14724304:T:A | rs112403360 | 14724304 | chr5:14811894:A:C | rs17252415 | 14811894 | -0.528 | 3.00E-04 | 87.6 | 1.17E-06 | 0.002 | 3.85E-04 | 0.397 |
| JAZF1 | 7 | chr7:28129126:GTCTT:G | rs1160871 | 28129126 | chr7:27868379:T:C | rs12700848 | 27868379 | 0.415 | 1.43E-04 | 260.7 | 4.41E-03 | 0.188 | 1.49E-02 | 0.337 |
| SEC61G | 7 | chr7:54873635:C:T | rs76928645 | 54873635 | chr7:55021758:A:T | rs12718937 | 55021758 | -0.383 | 1.70E-05 | 148.1 | 7.53E-03 | 0.383 | 2.44E-02 | 0.656 |
| SORL1 | 11 | chr11:121482368:T:G | rs74685827 | 121482368 | chr11:121367885:T:C | rs1560405 | 121367885 | 0.331 | 8.13E-04 | 114.5 | 5.39E-04 | 0.399 | 1.11E-05 | 0.025 |
| SORL1 | 11 | chr11:121564878:T:C | rs11218343 | 121564878 |  |  |  |  |  | 197.0 | 8.92E-07 | 0.002 | 3.93E-04 | 0.211 |
| FERMT2 | 14 | chr14:52924962:A:G | rs17125924 | 52924962 | chr14:52688191:A:G | rs71422098 | 52688191 | 0.521 | 2.06E-04 | 236.8 | 1.05E-03 | 0.038 | 8.92E-04 | 0.418 |
| ABCA7 | 19 | chr19:1050875:A:G | rs12151021 | 1050875 | chr19:1103523:A:G | rs8103283 | 1103523 | 0.497 | 8.18E-06 | 52.6 | 4.62E-03 | 0.081 | 1.92E-02 | 0.168 |
| ADAMTS1 | 21 | chr21:26775872:C:T | rs2830489 | 26775872 | chr21:27041769:C:G | rs77580019 | 27041769 | -0.858 | 1.63E-05 | 265.9 | 3.99E-03 | 0.14 | 5.62E-03 | 0.149 |
| NDUFAF6<sup>a</sup> | 8 | chr8:94983473:C:T | rs13276936 | 94983473 | chr8:95072607:C:A | rs2044899 | 95072607 | -0.4046 | 1.90E-05 | 89.1 | 2.42E-01 | 0.694 | 2.60E-01 | 0.766 |
| NDUFAF6<sup>b</sup> | 8 | chr8:94979792:C:T | rs10098778 | 94979792 |  |  |  |  |  | 92.8 | 2.12E-01 | 0.596 | 2.07E-01 | 0.635 |

**Supplementary Table 4.** Information of the SNPs included in the polygenic risk score extracted from Bellenguez et al. 2022 (EADB, StageII). Abbreviation: EADB, European Alzheimer’s and Dementia Biobank.

| ID | RS | Chromosome | Position (bp) | A1 (Effect allele) | A2 (Other Allele) | Beta | SE | P-value | A1 Frequency |
| --- | --- | --- | --- | --- | --- | --- | --- | --- | --- |
| chr1:109345810:T:C | rs141749679 | 1 | 109345810 | C | T | 0.3450 | 0.0938 | 2.36E-04 | 0.0038 |
| chr1:207577223:T:C | rs679515 | 1 | 207577223 | T | C | 0.1160 | 0.0151 | 1.51E-14 | 0.1877 |
| chr2:9558882:A:G | rs72777026 | 2 | 9558882 | G | A | 0.0550 | 0.0192 | 4.14E-03 | 0.1436 |
| chr2:37304796:T:C | rs17020490 | 2 | 37304796 | C | T | 0.0623 | 0.0169 | 2.33E-04 | 0.1446 |
| chr2:105749599:T:C | rs143080277 | 2 | 105749599 | C | T | 0.3677 | 0.1076 | 6.29E-04 | 0.0051 |
| chr2:127135234:C:T | rs6733839 | 2 | 127135234 | T | C | 0.1444 | 0.0126 | 2.06E-30 | 0.3891 |
| chr2:202878716:TC:T | rs139643391 | 2 | 202878716 | TC | T | 0.0699 | 0.0281 | 1.29E-02 | 0.8689 |
| chr2:233117202:G:C | rs10933431 | 2 | 233117202 | C | G | 0.0419 | 0.0148 | 4.70E-03 | 0.7657 |
| chr3:155069722:G:A | rs16824536 | 3 | 155069722 | G | A | 0.0861 | 0.0287 | 2.75E-03 | 0.9458 |
| chr3:155084189:A:G | rs61762319 | 3 | 155084189 | G | A | 0.1689 | 0.0456 | 2.14E-04 | 0.0263 |
| chr4:993555:G:T | rs3822030 | 4 | 993555 | T | G | 0.0454 | 0.0159 | 4.31E-03 | 0.5712 |
| chr4:11023507:C:T | rs6846529 | 4 | 11023507 | C | T | 0.0583 | 0.0140 | 3.16E-05 | 0.2826 |
| chr4:40197226:G:C | rs2245466 | 4 | 40197226 | G | C | 0.0448 | 0.0137 | 1.05E-03 | 0.3431 |
| chr5:14724304:T:A | rs112403360 | 5 | 14724304 | A | T | 0.1277 | 0.0314 | 4.63E-05 | 0.0727 |
| chr5:86927378:T:C | rs62374257 | 5 | 86927378 | C | T | 0.0571 | 0.0184 | 1.96E-03 | 0.2295 |
| chr5:151052827:C:T | rs871269 | 5 | 151052827 | C | T | 0.0445 | 0.0131 | 6.58E-04 | 0.6736 |
| chr5:180201150:G:A | rs113706587 | 5 | 180201150 | A | G | 0.0860 | 0.0197 | 1.30E-05 | 0.1103 |
| chr6:32615322:A:G | rs6605556 | 6 | 32615322 | A | G | 0.0662 | 0.0193 | 5.83E-04 | 0.8387 |
| chr6:41036354:G:A | rs10947943 | 6 | 41036354 | G | A | 0.0711 | 0.0170 | 2.98E-05 | 0.8580 |
| chr6:41161469:C:T | rs143332484 | 6 | 41161469 | T | C | 0.3664 | 0.0680 | 7.08E-08 | 0.0126 |
| chr6:41161514:C:T | rs75932628 | 6 | 41161514 | T | C | 0.8336 | 0.1247 | 2.34E-11 | 0.0031 |
| chr6:41181270:A:G | rs60755019 | 6 | 41181270 | G | A | 0.4145 | 0.2027 | 4.09E-02 | 0.0042 |
| chr6:47517390:C:T | rs7767350 | 6 | 47517390 | T | C | 0.0970 | 0.0139 | 2.95E-12 | 0.2709 |
| chr6:114291731:T:C | rs785129 | 6 | 114291731 | T | C | 0.0389 | 0.0127 | 2.11E-03 | 0.3502 |
| chr7:7817263:T:C | rs6943429 | 7 | 7817263 | T | C | 0.0496 | 0.0125 | 7.46E-05 | 0.4205 |
| chr7:8204382:T:C | rs10952097 | 7 | 8204382 | T | C | 0.0643 | 0.0234 | 5.95E-03 | 0.1136 |
| chr7:12229967:C:A | rs13237518 | 7 | 12229967 | C | A | 0.0530 | 0.0123 | 1.58E-05 | 0.5885 |
| chr7:28129126:GTCTT:G | rs1160871 | 7 | 28129126 | GTCTT | G | 0.0498 | 0.0229 | 2.96E-02 | 0.7777 |
| chr7:37844191:T:C | rs6966331 | 7 | 37844191 | C | T | 0.0542 | 0.0125 | 1.36E-05 | 0.6506 |
| chr7:54873635:C:T | rs76928645 | 7 | 54873635 | C | T | 0.0602 | 0.0198 | 2.32E-03 | 0.8967 |
| chr7:100334426:C:T | rs7384878 | 7 | 100334426 | T | C | 0.0816 | 0.0133 | 7.41E-10 | 0.6900 |
| chr7:143413669:G:A | rs11771145 | 7 | 143413669 | G | A | 0.0384 | 0.0126 | 2.29E-03 | 0.6524 |
| chr8:11844613:G:C | rs1065712 | 8 | 11844613 | C | G | 0.0549 | 0.0245 | 2.53E-02 | 0.0530 |
| chr8:27362470:C:T | rs73223431 | 8 | 27362470 | T | C | 0.0715 | 0.0125 | 1.17E-08 | 0.3694 |
| chr8:27607795:T:C | rs11787077 | 8 | 27607795 | C | T | 0.0888 | 0.0124 | 6.60E-13 | 0.6080 |
| chr8:144103704:G:A | rs34173062 | 8 | 144103704 | A | G | 0.1325 | 0.0300 | 1.02E-05 | 0.0814 |
| chr9:104903697:C:G | rs1800978 | 9 | 104903697 | G | C | 0.0438 | 0.0183 | 1.65E-02 | 0.1300 |
| chr10:11676714:A:G | rs7912495 | 10 | 11676714 | G | A | 0.0662 | 0.0121 | 5.02E-08 | 0.4619 |
| chr10:60025170:T:G | rs7068231 | 10 | 60025170 | G | T | 0.0547 | 0.0124 | 9.67E-06 | 0.5974 |
| chr10:80494228:C:T | rs6586028 | 10 | 80494228 | T | C | 0.0704 | 0.0150 | 2.51E-06 | 0.8036 |
| chr10:96266650:G:A | rs6584063 | 10 | 96266650 | A | G | 0.1238 | 0.0314 | 8.26E-05 | 0.9565 |
| chr10:122413396:A:G | rs7908662 | 10 | 122413396 | A | G | 0.0457 | 0.0122 | 1.71E-04 | 0.5326 |
| chr11:47370397:G:A | rs10437655 | 11 | 47370397 | A | G | 0.0498 | 0.0157 | 1.52E-03 | 0.3987 |
| chr11:60254475:G:A | rs1582763 | 11 | 60254475 | G | A | 0.1181 | 0.0128 | 2.87E-20 | 0.6290 |
| chr11:86157598:T:C | rs3851179 | 11 | 86157598 | C | T | 0.0947 | 0.0125 | 4.29E-14 | 0.6416 |
| chr11:121482368:T:G | rs74685827 | 11 | 121482368 | G | T | 0.1080 | 0.0516 | 3.65E-02 | 0.0186 |
| chr11:121564878:T:C | rs11218343 | 11 | 121564878 | T | C | 0.1961 | 0.0348 | 1.76E-08 | 0.9610 |
| chr12:113281983:T:C | rs6489896 | 12 | 113281983 | C | T | 0.0754 | 0.0201 | 1.78E-04 | 0.0764 |
| chr14:52924962:A:G | rs17125924 | 14 | 52924962 | G | A | 0.1067 | 0.0205 | 2.07E-07 | 0.0892 |
| chr14:92464917:G:A | rs7401792 | 14 | 92464917 | G | A | 0.0387 | 0.0125 | 1.91E-03 | 0.3709 |
| chr14:92472511:G:A | rs12590654 | 14 | 92472511 | G | A | 0.0654 | 0.0129 | 3.55E-07 | 0.6721 |
| chr14:106665591:G:A | rs10131280 | 14 | 106665591 | G | A | 0.0618 | 0.0179 | 5.42E-04 | 0.8675 |
| chr15:50701814:A:G | rs8025980 | 15 | 50701814 | A | G | 0.0593 | 0.0164 | 2.96E-04 | 0.6552 |
| chr15:58764824:T:A | rs602602 | 15 | 58764824 | T | A | 0.0530 | 0.0129 | 3.91E-05 | 0.7205 |
| chr15:63277703:C:T | rs117618017 | 15 | 63277703 | T | C | 0.0864 | 0.0198 | 1.21E-05 | 0.1439 |
| chr15:64131307:G:A | rs3848143 | 15 | 64131307 | G | A | 0.0650 | 0.0148 | 1.11E-05 | 0.2199 |
| chr15:78936857:A:G | rs12592898 | 15 | 78936857 | G | A | 0.0609 | 0.0183 | 8.69E-04 | 0.8673 |
| chr16:30010081:C:T | rs1140239 | 16 | 30010081 | C | T | 0.0459 | 0.0186 | 1.37E-02 | 0.6211 |
| chr16:31111250:C:T | rs889555 | 16 | 31111250 | C | T | 0.0392 | 0.0132 | 3.06E-03 | 0.7190 |
| chr16:70660097:C:A | rs4985556 | 16 | 70660097 | A | C | 0.0861 | 0.0197 | 1.21E-05 | 0.1148 |
| chr16:79574511:T:C | rs450674 | 16 | 79574511 | T | C | 0.0252 | 0.0125 | 4.31E-02 | 0.6272 |
| chr16:81739398:G:A | rs12446759 | 16 | 81739398 | A | G | 0.0375 | 0.0128 | 3.41E-03 | 0.5970 |
| chr16:86420604:T:A | rs16941239 | 16 | 86420604 | A | T | 0.1164 | 0.0359 | 1.20E-03 | 0.0288 |
| chr16:90103687:G:A | rs56407236 | 16 | 90103687 | A | G | 0.1025 | 0.0265 | 1.09E-04 | 0.0693 |
| chr17:1728046:TGAG:T | rs35048651 | 17 | 1728046 | T | TGAG | 0.1012 | 0.0220 | 4.11E-06 | 0.2137 |
| chr17:5233752:G:A | rs7225151 | 17 | 5233752 | A | G | 0.0461 | 0.0173 | 7.75E-03 | 0.1241 |
| chr17:18156140:G:A | rs2242595 | 17 | 18156140 | G | A | 0.0708 | 0.0175 | 5.16E-05 | 0.8883 |
| chr17:44352876:C:T | rs5848 | 17 | 44352876 | T | C | 0.0797 | 0.0132 | 1.46E-09 | 0.2886 |
| chr17:46779275:G:C | rs199515 | 17 | 46779275 | C | G | 0.0703 | 0.0168 | 2.78E-05 | 0.7806 |
| chr17:58332680:A:G | rs2526377 | 17 | 58332680 | A | G | 0.0547 | 0.0121 | 6.71E-06 | 0.5551 |
| chr17:63471557:C:T | rs4277405 | 17 | 63471557 | T | C | 0.0538 | 0.0125 | 1.60E-05 | 0.6163 |
| chr19:1050875:A:G | rs12151021 | 19 | 1050875 | A | G | 0.0833 | 0.0139 | 2.36E-09 | 0.3357 |
| chr19:1854254:G:GC | rs149080927 | 19 | 1854254 | G | GC | 0.0419 | 0.0184 | 2.30E-02 | 0.4802 |
| chr19:49950060:C:T | rs9304690 | 19 | 49950060 | T | C | 0.0549 | 0.0146 | 1.66E-04 | 0.2398 |
| chr19:54267597:C:T | rs587709 | 19 | 54267597 | C | T | 0.0509 | 0.0138 | 2.21E-04 | 0.3251 |
| chr20:413334:A:G | rs1358782 | 20 | 413334 | G | A | 0.0457 | 0.0150 | 2.36E-03 | 0.7540 |
| chr20:56423488:A:G | rs6014724 | 20 | 56423488 | A | G | 0.1132 | 0.0236 | 1.56E-06 | 0.9102 |
| chr20:63743088:T:C | rs6742 | 20 | 63743088 | C | T | 0.0604 | 0.0164 | 2.22E-04 | 0.7790 |
| chr21:26101558:C:T | rs2154481 | 21 | 26101558 | T | C | 0.0445 | 0.0120 | 2.15E-04 | 0.5236 |
| chr21:26775872:C:T | rs2830489 | 21 | 26775872 | C | T | 0.0337 | 0.0135 | 1.28E-02 | 0.7191 |

**Supplementary Table 5.**
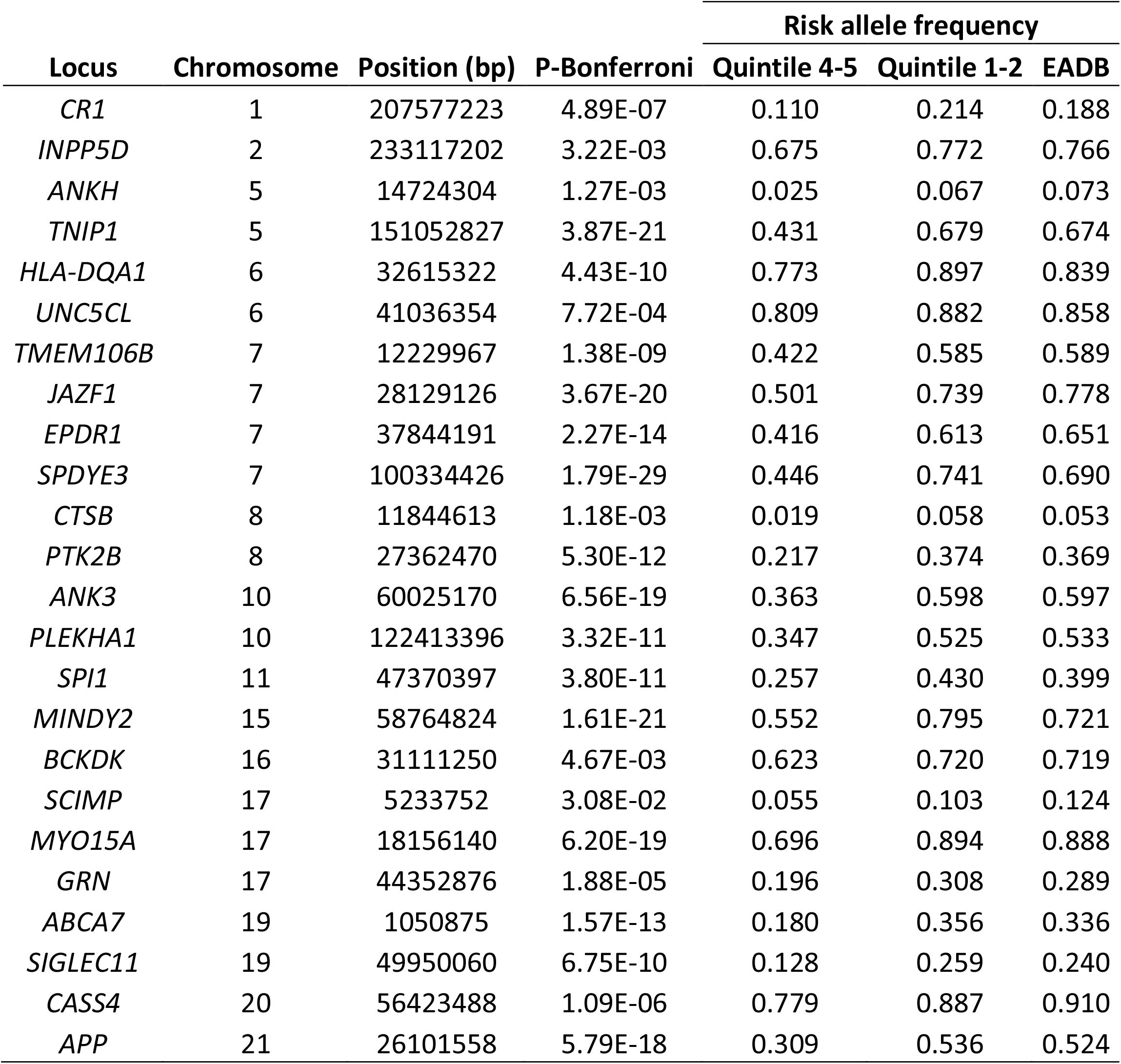
Risk alleles less frequent in Native American ancestry. Abbreviation: EADB, European Alzheimer’s and Dementia Biobank.

**Supplementary Table 6.** Risk alleles more frequent in Native American ancestry. Abbreviation: EADB, European Alzheimer’s and Dementia Biobank.

| Locus | Chromosome | Position (bp) | P Bonferroni | Risk allele frequency |  |  |
| --- | --- | --- | --- | --- | --- | --- |
|  |  |  |  | Quintile 4-5 | Quintile 1-2 | EADB |
| PRKD3 | 2 | 37304796 | 1.39E-35 | 0.460 | 0.140 | 0.145 |
| BIN1 | 2 | 127135234 | 1.46E-08 | 0.509 | 0.362 | 0.389 |
| RHOH | 4 | 40197226 | 1.13E-03 | 0.448 | 0.345 | 0.343 |
| TREML2 | 6 | 41181270 | 7.58E-03 | 0.037 | 0.005 | 0.004 |
| HS3STS | 6 | 114291731 | 5.55E-06 | 0.479 | 0.358 | 0.350 |
| SNX1 | 15 | 64131307 | 1.14E-12 | 0.397 | 0.228 | 0.220 |
| CTSH | 15 | 78936857 | 4.99E-02 | 0.933 | 0.889 | 0.867 |
| DOC2A | 16 | 30010081 | 3.93E-05 | 0.760 | 0.656 | 0.621 |
| MAPT | 17 | 46779275 | 1.21E-08 | 0.871 | 0.754 | 0.781 |
| ACE | 17 | 63471557 | 2.23E-10 | 0.747 | 0.588 | 0.616 |
| KLF16 | 19 | 1854254 | 5.31E-12 | 0.663 | 0.479 | 0.480 |
| RBCK1 | 20 | 413334 | 1.06E-16 | 0.866 | 0.679 | 0.754 |
| SLC2A4RG | 20 | 63743088 | 3.75E-08 | 0.902 | 0.794 | 0.779 |
| ADAMTS1 | 21 | 26775872 | 3.01E-12 | 0.855 | 0.711 | 0.719 |

